# Characterisation of the blood RNA host response underpinning severity in COVID-19 patients

**DOI:** 10.1101/2021.09.16.21263170

**Authors:** Heather Jackson, Irene Rivero Calle, Claire Broderick, Dominic Habgood-Coote, Giselle d’Souza, Samuel Nichols, Jose Gómez-Rial, Carmen Rivero-Velasco, Nuria Rodríguez-Núñez, Gema Barbeito-Castiñeiras, Hugo Pérez-Freixo, Manuel Barreiro-de Acosta, Aubrey J. Cunnington, Jethro A. Herberg, Victoria J. Wright, Alberto Gómez-Carballa, Antonio Salas, Mike Levin, Federico Martinon-Torres, Myrsini Kaforou, PERFORM consortium and GEN-COVID study group

## Abstract

Infection with SARS-CoV-2 has highly variable clinical manifestations, ranging from asymptomatic infection through to life-threatening disease. Host whole blood transcriptomics can offer unique insights into the biological processes underpinning infection and disease, as well as severity. We performed whole blood RNA Sequencing of individuals with varying degrees of COVID-19 severity. We used differential expression analysis and pathway enrichment analysis to explore how the blood transcriptome differs between individuals with mild, moderate, and severe COVID-19, performing pairwise comparisons between groups. Increasing COVID-19 severity was characterised by an abundance of inflammatory immune response genes and pathways, including many related to neutrophils and macrophages, in addition to an upregulation of immunoglobulin genes. Our insights into COVID-19 severity reveal the role of immune dysregulation in the progression to severe disease and highlight the need for further research exploring the interplay between SARS-CoV-2 and the inflammatory immune response.

## 1. Introduction

Since severe acute respiratory syndrome coronavirus 2 (SARS-CoV-2), the causative agent of coronavirus disease 19 (COVID-19), emerged in Wuhan (China) in late 2019, it has caused considerable morbidity and mortality, in addition to major impacts on global health and economic systems. Manifestation of SARS-CoV-2 infection is highly heterogeneous, with some individuals remaining asymptomatic and others developing COVID-19, which can range from mild flu-like symptoms to severe life-threatening disease requiring mechanical ventilation and intensive care, and even death ^1^. Most SARS-CoV-2 infections lead to mild symptoms or no symptoms at all, allowing cases to remain undetected and thus facilitating its spread throughout populations ^1,2^. It was recognised early during the pandemic that age and pre-existing health conditions, such as obesity and diabetes, are amongst the main risk factors for developing severe COVID-19 ^3,4^. Dysfunctional immune responses to SARS-CoV-2, specifically impaired type I interferon responses in conjunction with an exacerbated inflammatory response, have been associated with progression to severe COVID-19 ^5-7^. An imbalanced immune response leads to the development of a ‘cytokine storm’ which causes lung inflammation, septic shock, and multi-organ failure ^5,7,8^.

Identification of improved treatments and prevention of severe COVID-19 requires better understanding of the underlying immune and inflammatory processes that distinguish severe disease from mild illness. Host whole blood transcriptomic profiling of patients with infectious and inflammatory conditions has been extensively used for understanding infectious disease dynamics, from the identification of accurate biomarkers of infection ^9-11^ to gaining insights into variations in the host response to different pathogens and severity of disease ^12,13^. Targeted ^6,14^ and untargeted ^15-18^ transcriptomic profiling of whole blood from SARS-CoV-2-positive hosts has already been undertaken. To the best of our knowledge, severity of COVID-19 has not yet been explored in whole blood with non-hospitalised SARS-CoV-2-positive cases included as a comparator group, nor has it been explored across a range of severities. The study of transcriptomic profiles from individuals with varying severities will be an essential tool for improving our understanding of the course of disease resulting from SARS-CoV-2 infection.

We have analysed the whole blood transcriptomes obtained from individuals with different levels of COVID-19 severity to explore how the host blood transcriptome changes with increasing COVID-19 severity, aiming to identify the key biological processes and genes underpinning differences in severity.

## 2. Methods

### 2.1. Study design and clinical cohort

Adult patients with COVID-19 were recruited through the GEN-COVID study group (www.gencovid.eu) at Hospital Clínico Universitario de Santiago de Compostela (Galicia, Spain) between March 2020 and May 2020. COVID-19 was defined according to the current national guidelines in Spain (https://www.mscbs.gob.es/profesionales/saludPublica/ccayes/alertasActual/nCov/documentos.htm).

Subjects granted informed consent for their participation in the study. If this was not possible at the time of sampling, deferred consent was allowed, and subjects were approached for consent at the earliest appropriate opportunity. Subjects who did not agree to participate in the study were excluded. GEN-COVID Study was approved by the Ethics Committee of Galicia by fast-track procedure on 18^th^ March 2020 (CEIC Galicia, reg 2020/178).

Patients with COVID-19 were categorised as having mild, moderate, or severe disease. Mild patients were those who were always outpatients; emergency department attendance was the ceiling of care, and they were not admitted to hospital (WHO score 1-2). Moderate patients were those admitted to hospital, for whom ward-based therapy was the ceiling of care with supportive care limited to oxygen delivery; no intensive care unit (ICU) admission (WHO score 3-4). Severe patients were those who were admitted to ICU at any time throughout the course of their disease (WHO score 5-7). Supportive care included high flow oxygen (>16 litres/ minute), non-invasive ventilation (NIV), invasive ventilation, inotropic support, renal replacement therapy, and extracorporeal membrane oxygenation (ECMO). The severe category also included those patients who died (WHO score 8), in the emergency department, ward or ICU.

### 2.2 RNA isolation and quantification

Whole blood was collected at the time of recruitment into PAXgene blood RNA tubes (PreAnalytiX), frozen, and total RNA (including RNA > 18 nucleotides) was isolated according to the manufacturer’s instructions (Qiagen). RNA samples were stored at −80°C, before undergoing an additional DNAse treatment using an RNA clean & concentrator kit (Zymo Research) prior to sequencing at The Wellcome Centre for Human Genetics in Oxford, UK. Material was quantified using RiboGreen (Invitrogen) on the FLUOstar OPTIMA plate reader (BMG Labtech) and the size profile and integrity analysed on the 2200 TapeStation (Agilent, RNA ScreenTape). Input material was normalised and strand specific library preparation was completed using NEBNext® Ultra− II mRNA kit (NEB) and NEB rRNA/globin depletion probes following manufacturer’s instructions. Libraries were on a Tetrad (Bio-Rad) using in-house unique dual indexing primers (based on ^19^). Individual libraries were normalised using Qubit and pooled together. The pooled library was diluted to ∼10 nM for storage and denatured and further diluted prior to loading on the sequencer. Paired end sequencing was performed using a Novaseq6000 platform at 150 paired end configuration. The RNA-Seq analysis pipeline consisted of quality control using FastQC ^20^, MultiQC ^21^ and annotations modified with BEDTools ^22^, alignment and read counting using STAR ^23^, SAMtools ^24^, FeatureCounts ^25^ and version 89 ensembl GCh38 genome and annotation ^26^.

### 2.3. Statistical analysis

All statistical analyses were performed using the statistical software R (R version 4.0.3) ^27^. Normalised counts were calculated for each gene using DESeq2 (V1.30.0) ^28^ and default parameters were used. Normalised genes with fewer than three samples with a normalised read count of at least 20 were considered lowly expressed and were removed, leaving a total of 20,536 genes. Principal component analysis (PCA) was performed on the normalised counts.

Immune cell level measurements were not available for all individuals included in the analysis. Therefore, cell-type fractions were estimated from the bulk host transcriptome data using the CIBERSORTx algorithm ^29^. The estimated fractions were compared between the three COVID-19 patient groups (mild, moderate, and severe) and statistical significance was evaluated using a two-sided Mann-Whitney-Wilcoxon test with Bonferroni correction to correct for the multiple tests (3 test per cell type: moderate *vs*. mild; severe *vs*. mild; severe *vs*. moderate). Adjusted p-values < 0.05 were considered significant.

Prior to exploring the impact of COVID-19 severity on the transcriptome, the effect of immunomodulatory treatment on the transcriptome was assessed. Patients with moderate COVID-19 who were not receiving steroids at the time of sampling were contrasted against patients with moderate COVID-19 who were receiving steroids at the time of sampling, whilst accounting for age and sex in the model. Steroids were chosen as the immunomodulatory treatment to explore due to adequate sample size in comparison to other immunomodulatory treatment groups such as tocilizumab. Patients receiving tocilizumab or interferon therapies were excluded from this comparison. Treatments including antibiotics, antivirals and antimalarials were allowed.

DESeq2 ^28^ was used for differential expression analysis of COVID-19 severity groups. We assessed transcriptomic differences with increasing COVID-19 severity by carrying out pairwise comparisons between each severity group (i.e., moderate *vs*. mild; severe *vs*. mild; and severe *vs*. moderate). We used two different model designs for each comparison. First, the models included immunomodulatory treatment status, sex, age, and severity. This model design aimed to account for transcriptomic differences induced by immunomodulatory treatments by including variables representing whether the patients received tocilizumab, steroids, or interferon treatment, in addition to sex, age and severity. The second model design included *in silico* immune cell proportion estimates, sex, age, and severity. This design accounted for transcriptomic differences induced by different proportions of immune cells, and it included the immune cell fractions described below as well as sex, age, and severity. The immune cell proportions accounted for included: monocytes, neutrophils, B cells (the sum of naïve and memory B cells and plasma cell proportions), CD4 T cells, (the sum of the proportions of naïve CD4 T cells, resting and activated memory CD4 T cells, follicular helper T cells and regulatory T cells), CD8 T cells and natural killer (NK) cells (the sum of resting and activated NK cell proportions).

Adjusted p-values were calculated using the Benjamini-Hochberg (B-H) procedure ^30^. The log_2_ fold-changes (LFC) and adjusted p-values of all genes were visualised using volcano plots. Concordance and discordance resulting from different model designs were visualised using cross plots. Genes with an adjusted p-value < 0.05 were considered significantly differentially expressed (SDE). The lists of SDE genes were subjected to pathway analysis using Ingenuity Pathway Analysis (IPA; QIAGEN Inc., https://www.qiagenbioinformatics.com/products/ingenuity-pathway-analysis). IPA was selected because it can predict directionality of pathways through knowledge of molecular functions, and it returns a *z*-score for each predicted pathway. Positive and negative *z*-scores indicate that the pathway is upregulated or downregulated, respectively, in the group of interest compared against the reference group.

Severity was also explored as an additive variable (mild = 0; moderate = 1; severe = 2), with full methods described in the supplementary text.

## 3. Data availability

Gene counts and sample metadata are available on ArrayExpress under the accession E-MTAB-10926. Code used in the analyses can be found at: https://github.com/PIDBG/sarscov2_transcriptomics/tree/main/severity_analysis.

## 4. Results

A schematic showing an overview of the patients, analysis, and key findings is shown in Figure 1.

**Figure 1.**
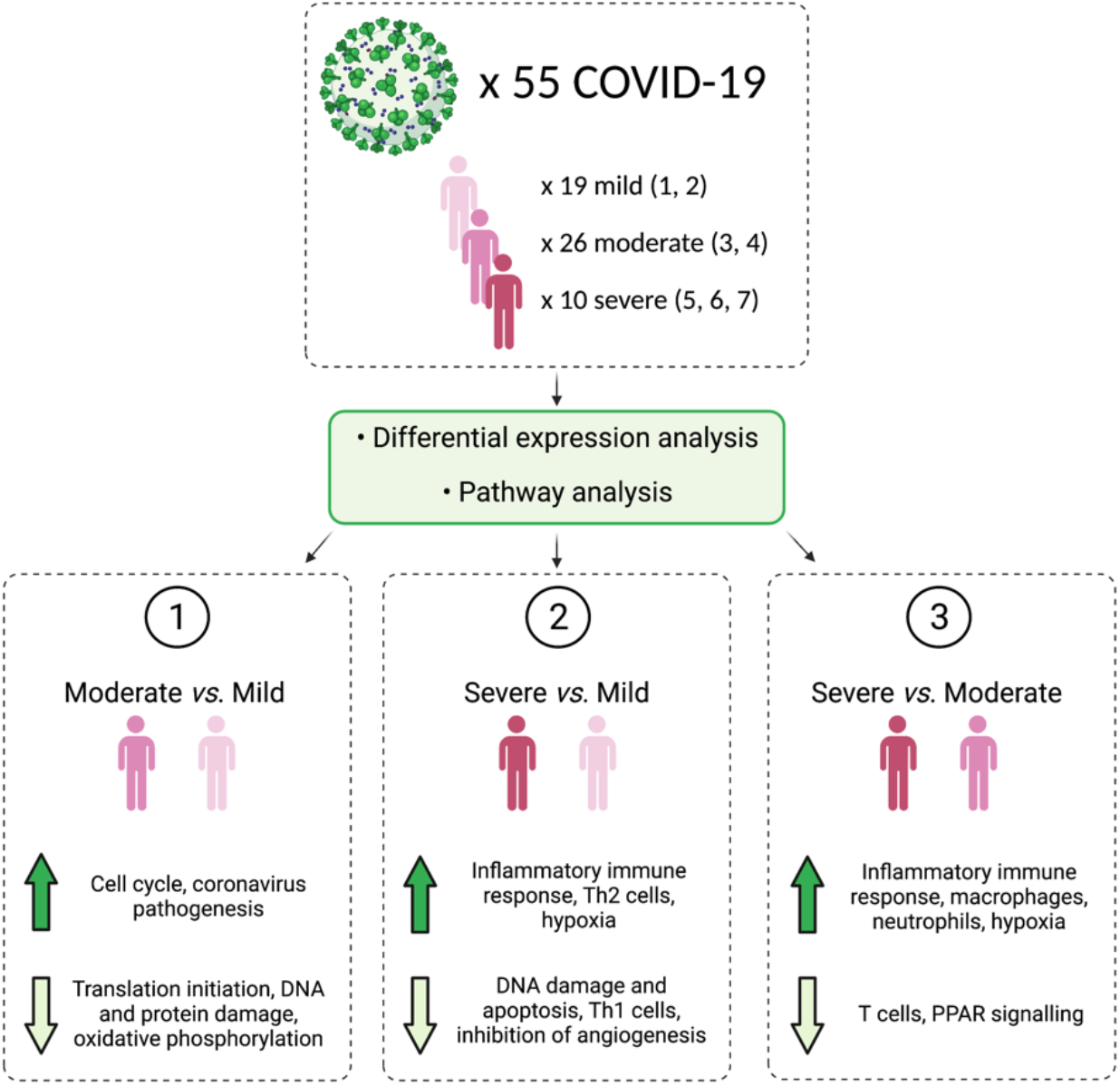
A schematic summarising the number of patients analysed, the main analysis steps, and the key findings. The numbers in brackets following mild, moderate, and severe are the WHO severity scores that make up these classifications. Figure made with BioRender (https://biorender.com/).

### 4.1. Clinical Description of patients

Whole blood transcriptomic profiling through RNA Sequencing (RNA-Seq) was performed on 65 samples from patients recruited through the GEN-COVID study. Samples from patients in whom a pathogen in addition to SARS-CoV-2 was isolated less than 5 days before or 10 days after the research blood sample were not selected (*n*=10), as the aim was to identify gene expression changes in blood that reflect COVID-19 disease and not coinfections. Of the 55 remaining COVID-19 patients, 19, 26 and 10 patients had mild, moderate, and severe disease, respectively. In all but two cases, the severity categorisation of the patient matched their level of supportive care at the time the research blood sample. For these two patients, the decision to transfer them to ICU was made within 36 hours of the sample extraction, therefore they were classified as severe in our analyses.

Characteristics of the 55 COVID-19 patients are summarised in Table 1 and their WHO Severity Classification Score and how it relates to the severity groups used here is detailed in Table 2. 52.7% (*n*=29) were female and 98% (*n*=54) were South European. The median age was 55 years. 78.2% (*n*=43) of the patients reported comorbidities with endocrine conditions (most frequently diabetes) as the most common comorbidities (45.4%; *n*=25) followed by obesity (32.7%; *n*=18) and hypertension (18.2%; *n*=10). The duration of symptoms before presentation to Emergency Department (ED) was a median of 13 days. Main presenting symptoms were respiratory (80.3%; *n*=45), fever (74.5%; *n*=41) and musculoskeletal (56.4%; *n*=31). All patients had PCR-confirmed SARS-CoV-2 infection and 96.4% (*n*=53) were community acquired.

**Table 1.** Clinical characteristics of the patients included in the analysis (*n*=55) stratified by COVID-19 severity. The section ‘Supportive care required information’ refers to the care required at sampling whilst ‘Maximum level of care/outcome information’ refers to the maximum level of care required for the patients after sampling. High-flow oxygen is oxygen therapy between 16L/min - 60L/min.

| | Mild ($n=19$) | Moderate ($n=26$) | Severe ($n=10$) |
| --- | --- | --- | --- |
| **General demographics** |  |  |  |
| Sex (female) | 14 (73.7%) | 12 (46.2%) | 3 (30%) |
| Age (median) | 37 (32.5-42.5) | 69 (56.5-76.8) | 65 (54.3-70) |
| <50 years | 19 (100%) | 5 (19.2%) | 0 (0%) |
| 50-70 years | 0 (0%) | 9 (34.6%) | 6 (60%) |
| >70 years | 0 (0%) | 12 (46.2%) | 4 (40%) |
| Ethnicity (South European) | 19 (100%) | 25 (96.2%) | 10 (100%) |
| **Comorbidities** |  |  |  |
| Endocrine | 4 (21%) | 14 (53.8 %) | 7 (70%) |
| Smoking | 1 (5.3%) | 4 (15.4%) | 5 (50%) |
| Malignancy | 0 (0%) | 0 (0%) | 1 (10%) |
| Pulmonary | 0 (0%) | 8 (30.8%) | 1 (10%) |
| Gastrointestinal | 0 (0%) | 5 (19.2%) | 1 (10%) |
| Neurological | 3 (15.8%) | 2 (7.7%) | 0 (0%) |
| Cardiac | 0 (0%) | 7 (26.9%) | 0 (0%) |
| Hypertension | 1 (5.3%) | 6 (23.1%) | 3 (30%) |
| Immunosuppressed | 0 (0%) | 1 (3.8%) | 1 (10%) |
| Hematology | 0 (0%) | 0 (0%) | 0 (0%) |
| Allergic/autoimmune/inflammatory | 2 (10.5%) | 0 (0%) | 0 (0%) |
| Genetic | 0 (0%) | 1 (3.8%) | 0 (0%) |
| Organ transplant | 0 (0%) | 0 (0%) | 1 (10%) |
| Obesity | 1 (5.3%) | 12 (46.2%) | 5 (50%) |
| Pregnant | 1 (5.3%) | 0 (0%) | 0 (0%) |
| Other | 0 (0%) | 0 (0%) | 0 (0%) |
| **Admission timelines** |  |  |  |
| Days since symptom onset (median, IQR) | 11 (9.25-16.75) | 15 (11-17) | 10.5 (9-13.5) |
| **Presenting symptoms** |  |  |  |
| Fever | 9 (47.4%) | 22 (84.6%) | 10 (100%) |
| Respiratory | 12 (63.2%) | 24 (92.3%) | 9 (90%) |
| Cardiac | 0 (0%) | 1 (3.8%) | 0 (0%) |
| Gastrointestinal | 2 (10.5%) | 9 (34.6%) | 4 (40%) |
| Musculoskeletal | 9 (47.4%) | 16 (61.5%) | 6 (60%) |
| Rash | 0 (0%) | 0 (0%) | 0 (0%) |
| Sensory (ageusia, anosmia) | 9 (47.4%) | 8 (30.8%) | 1 (10%) |
| Headache | 10 (52.6%) | 4 (15.4%) | 0 (0%) |
| Shock | 0 (0%) | 0 (0%) | 0 (0%) |
| Ill appearance | 0 (0%) | 2 (7.7%) | 1 (10%) |
| **Investigations** |  |  |  |
| White cells ($10^9/L$) – mean (IQR) | Unknown | 7 (4.6-6.9) | 7.5 (5.2-10) |
| Neutrophils ($10^9/L$) – mean (IQR) | Unknown | 4.5 (2.6-4.7) | 6.9 (4.5-9.1) |
| Lymphocytes ($10^9/L$) – mean (IQR) | Unknown | 1.6 (1.1-2.1) | 0.65 (0.5-0.7) |
| Monocytes ($10^9/L$) – mean (IQR) | Unknown | 0.41 (0.32-0.51) | 0.26 (0.14-0.38) |
| Platelets ($10^9/L$) – mean (IQR) | Unknown | 317.1 (242.5-381) | 224.9 (128.8-340) |
| Fibrinogen (mg/dL)- mean (IQR) | Unknown | 475.6 (476.5-500) | 444.6 (379.5-511.5) |
| D-dimer (ng/mL)- mean (IQR) | Unknown | 1694.7 (526-1132) | 4122 (708-6267) |
| CRP (mg/L)- mean (IQR) | Unknown | 34.4 (16.2-36.4) | 81.8 (29.1-127.8) |
| Ferritin (µg/L)- mean (IQR) | Unknown | 664.5 (255.8-734) | 1604 (1196-1817) |
| **SARS-CoV-2 infection acquisition (if known)** |  |  |  |
| Community acquired infection | 18 (94.7%) | 25 (96.2%) | 10 (100%) |
| Hospital acquired infection | 0 (0%) | 0 (0%) | 0 (0%) |
| **Treatment** |  |  |  |
| Antibiotic | 0 (0%) | 26 (100%) | 10 (100%) |
| Antiviral (lopinavir-ritonavir) | 0 (0%) | 22 (84.6%) | 0 (0%) |
| Antimalarial (hydroxychloroquine) | 0 (0%) | 25 (96.2%) | 10 (100%) |
| Steroids | 0 (0%) | 7 (26.9%) | 5 (50%) |
| Monoclonal antibody (tocilizumab, infliximab, vedolizumab) | 0 (0%) | 1 (3.8%) | 4 (40%) |
| **Supportive care required at time of sampling** |  |  |  |
| Oxygen | 0 (0%) | 15 (57.7%) | 0 (0%) |
| High-flow oxygen | 0 (0%) | 0 (0%) | 5 (50%) |
| Non-invasive ventilation | 0 (0%) | 0 (0%) | 1 (10%) |
| Invasive ventilation | 0 (0%) | 0 (0%) | 4 (40%) |
| Inotropes | 0 (0%) | 0 (0%) | 2 (20%) |
| ECMO | 0 (0%) | 0 (0%) | 0 (0%) |
| Renal replacement | 0 (0%) | 0 (0%) | 0 (0%) |
| Hemofiltration | 0 (0%) | 0 (0%) | 0 (0%) |
| **Maximum level of care/outcome** |  |  |  |
| Ambulatory | 19 (100%) | 0 (0%) | 0 (0%) |
| Admitted to ward | 0 (0%) | 26 (100%) | 0 (0%) |
| Admitted to ICU | 0 (0%) | 0 (0%) | 10 (100%) |
| Died | 0 (0%) | 0 (0%) | 0 (0%) |

**Table 2.** WHO Severity Classification Score for the COVID-19 patients included in the analysis. This is the patients’ final severity score rather than their severity at the time of sampling (as shown in table 1).

| WHO Severity Classification | Mild COVID-19 (n=19) | Moderate COVID-19 (n=26) | Severe COVID-19 (n=10) |
| --- | --- | --- | --- |
| Uninfected | 0 (0%) | 0 (0%) | 0 (0%) |
| **Ambulatory** |  |  |  |
| 1 - No limitation of activities | 19 (100%) | 0 (0%) | 0 (0%) |
| 2 - Limitation of activities | 0 (0%) | 0 (0%) | 0 (0%) |
| **Hospitalised, Mild Disease** |  |  |  |
| 3 - Hospitalised, no oxygen therapy | 0 (0%) | 5 (19.3%) | 0 (0%) |
| 4 - Oxygen by mask or nasal prongs | 0 (0%) | 21 (80.8%) | 0 (0%) |
| **Hospitalised, Severe Disease** |  |  |  |
| 5 - Non-invasive ventilation or high-flow oxygen | 0 (0%) | 0 (0%) | 4 (40%) |
| 6 - Intubation and mechanical ventilation | 0 (0%) | 0 (0%) | 3 (60%) |
| 7 - Ventilation + additional organ support- pressors, renal replacement therapy, ECMO | 0 (0%) | 0 (0%) | 3 (60%) |
| 8 - Death | 0 (0%) | 0 (0%) | 0 (0%) |

A combined triple therapy (antibiotic + antiviral [lopinavir-ritonavir] + antimalarial [hydroxychloroquine]) was the most common treatment administered to patients (*n*=30; 54.5%). 21.8% (*n*=12) received steroids and 9% (*n*=5) received tocilizumab during their disease. 54.5% (*n*=30) required oxygen at some point throughout the entire course of their disease, 7% (*n*=4) required invasive ventilation and 3.6% (*n*=2) required inotropes. Regarding outcomes, 34.5% (*n*=19) were ambulatory patients, 47% (*n*=26) were admitted to the ward, 18.2% (*n*=10) were admitted to ICU, and none died. Supplementary Fig. 1 shows that whilst the samples are clearly stratified by severity in PC1, there is confounding between severity and sex/age, a pattern which is also clear in Table 1.

### 4.2. In silico immune cell proportion estimates

As COVID-19 is associated with changes in blood cell proportions, particularly between approximately days 4-14 ^31-33^, that are more prominent among critically ill patients with COVID-19 ^34,35^, we assessed the levels of the cell-type proportions estimated *in-silico* from the RNA-Seq count data (Supplementary Fig. 2). Two-sided Mann-Whitney-Wilcoxon tests with Bonferroni correction were used to test for significance with adjusted p-values < 0.05 considered significant. CD4 T cell proportions were significantly different between all three pairwise severity comparisons with levels decreasing with increasing severity (moderate *vs*. mild *p*-value=5.5×10^−04^; severe *vs*. mild *p*-value=1.4×10^−05^; severe *vs*. moderate *p*-value=4.2×10^−03^). The proportions of CD8 T cells, neutrophils, monocytes and NK cells were significantly different between severe *vs*. mild COVID-19 (CD8 T cell *p*-value=6.8×10^−04^, neutrophil *p*-value=1.1×10^−04^, monocyte *p*-value=6.3×10^−03^, NK cell *p*-value=5.7×10^−03^) and severe *vs*. moderate COVID-19 (CD8 T cell *p*-value=0.014, neutrophil *p*-value=2.3×10^−04^, monocyte *p*-value=2.8×10^−03^, NK cell *p*-value=8.8×10^−03^). CD8 T cell, monocyte and NK cell proportions decreased between severe COVID-19 and mild/moderate COVID-19 whilst neutrophil proportions increased between moderate and severe cases. Since many mild patients were missing clinical cell counts, the *in silico* immune cell proportion estimates were used in downstream analyses in lieu of clinical cell counts.

### 4.3. The effect of immunomodulatory treatment on COVID-19 patients’ blood transcriptome

Patients received various types of clinical interventions and treatments (Table 1), including immunomodulatory therapies. To assess the impact of immunomodulatory therapies on the transcriptome of individuals with COVID-19, we identified genes SDE between patients with moderate COVID-19 who did not receive steroids (*n*=19) and patients with moderate COVID-19 who did receive steroids, excluding those who also received monoclonal antibody therapy (*n*=6). Individuals who received steroids in combination with antibiotics, antivirals and antimalarials were included in the comparisons. 556 genes were SDE in patients who received steroids *vs*. those who did not (B-H *p*-value < 0.05), of which 253 genes were over-expressed with steroid administration and 303 were under-expressed. No significant pathways were identified by IPA.

### 4.4. Differential gene expression analysis of COVID-19 severity

Pairwise comparisons were made between the three severity groups (mild, moderate and severe) to identify transcriptomic differences with increasing severity. In the moderate *vs*. mild COVID-19 comparison, there was greater concordance between the models accounting for immunomodulatory treatment and immune cell proportions as the number of genes identified as SDE in both models was higher than for the severe *vs*. mild and the severe *vs*. moderate comparisons (Fig. 2).

**Figure 2.**
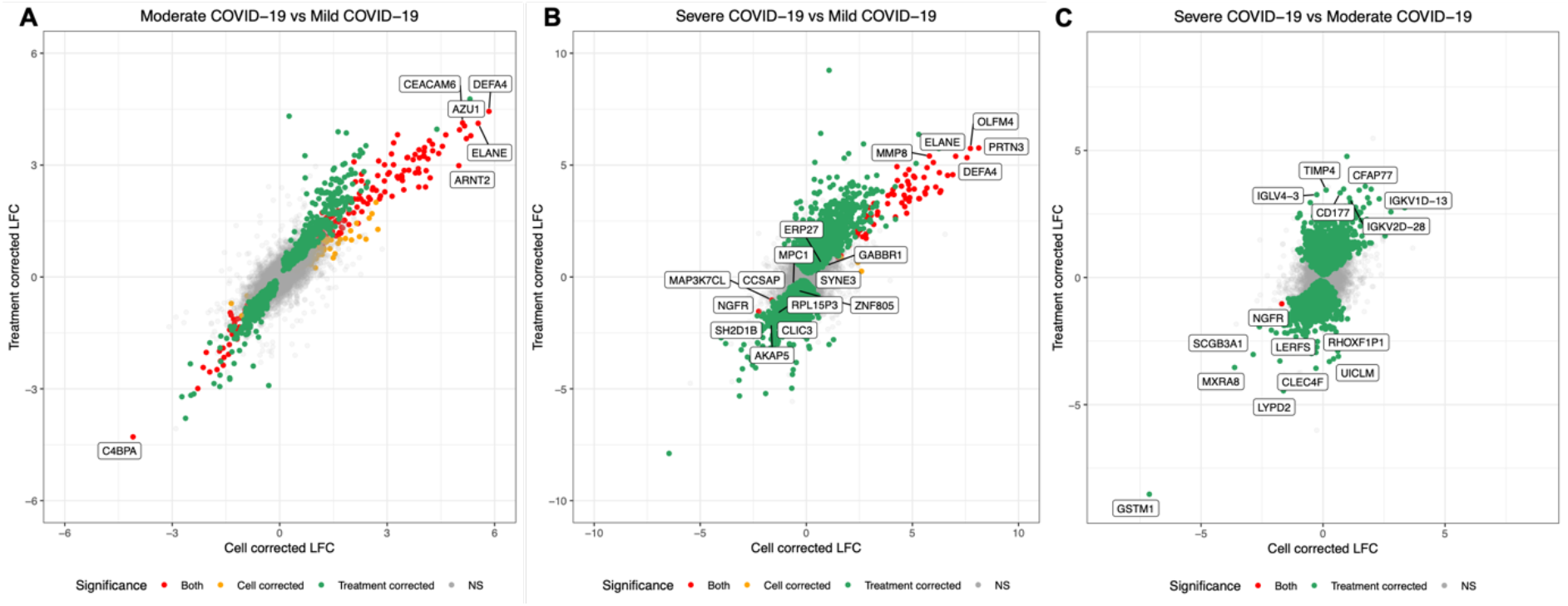
Cross plots showing the log_2_ fold change (LFC) values of genes for pairwise comparisons between three severity groups (**A**: moderate *vs*. mild; **B**: severe *vs*. mild; **C**: severe *vs*. moderate). The plots show how LFC values differ according to whether immune cell proportions (x-axis) or immunomodulatory treatments (y-axis) were included in the models. Red points are genes that were SDE in both models, whilst orange and green points are genes SDE in the cell correction and treatment correction models, respectively. NS = not significant.

#### Moderate COVID-19 vs. Mild COVID-19

1,547 genes were SDE between moderate (*n*=26) and mild (*n*=19) patients whilst accounting for immunomodulatory treatment, with 603 and 944 genes over- and under-expressed with increasing severity respectively. When these genes were subjected to pathway analysis, EIF2 Signalling was the most significant pathway reduced in moderate cases compared to mild cases (*z*-score=-5.778, B-H *p*-value=5.012×10^−29^) and Regulation of eIF4 and p70S6K Signalling (*z-*score=-1, B-H *p*-value=1.000×10^−13^) were also found to be significantly enriched (Supplementary Table 1).

The Coronavirus Pathogenesis pathway was upregulated in moderate COVID-19 (*z-*score=2.744, B-H *p*-value=5.248×10^−07^) in addition to various pathways related to the cell cycle including Mitotic Roles of Polo-Like Kinase (*z*-score=0.816, B-H *p*-value=2.754×10^−05^), Cyclins and Cell Cycle Regulation (*z*-score=2.714, B-H *p*-value=4.266×10^−02^) and Cell Cycle Control of Chromosomal Replication (*z*-score=0.632; B-H *p*-value=2.042×10^−02^).

PAK signalling (*z*-score=-2.53; B-H *p*-value=4.266×10^−02^) and mTOR signalling (*z*-score=-0.632; B-H *p*-value=4.467×10^−08^) pathways were both found to be downregulated in moderate COVID-19. Two pathways related to aberrant protein production and DNA damage were identified as downregulated in moderate COVID-19 whilst accounting for treatment: Unfolded Protein Response (*z*-score=-1; B-H *p*-value=1.514×10^−03^); and Role of CHK Proteins in Cell Cycle Checkpoint Control (*z*-score=-2.828; B-H *p*-value=7.943×10^−03^). Oxidative phosphorylation was also identified by IPA as downregulated in moderate COVID-19 (*z*-score=-4.243; B-H *p*-value=1.514×10^−03^).

488 genes were SDE between moderate and mild COVID-19 whilst accounting for immune cell proportions, with 222 and 266 genes over- and under-expressed respectively with increasing severity. 389 genes were SDE between moderate and mild COVID-19 irrespective of the model design (Fig. 2). When immune cell proportions were included in the model, IPA identified two significant pathways (EIF2 Signalling: *z*-score=-2.53, B-H *p* value=1.288×10^−03^; Regulation of eIF4 and p70S6K Signalling: B-H *p*-value=1.950×10^−02^; indeterminate z-score).

#### Severe COVID-19 vs. Mild COVID-19

7,343 genes were SDE between severe (*n*=10) and mild COVID-19 (*n*=19) whilst accounting for immunomodulatory treatment with 3,329 and 4,014 genes over- and under-expressed with increasing severity, respectively. 94 genes were SDE between severe and mild COVID-19 whilst accounting for immune cell proportions with 81 and 13 genes over- and under-expressed with increasing severity, respectively. 87 genes were SDE between severe and mild COVID-19 irrespective of the model design (Fig. 2B).

The pathways upregulated in severe COVID-19 were dominated by those related to the immune response, notably the inflammatory immune response (Supplementary Table 2). For example, TREM1 Signalling (*z*-score=3.157; B-H *p*-value=3.162×10^−02^), STAT3 Pathway (*z*-score=0.949; B-H *p*- value=7.762×10^−03^) and IL-22 signalling (*z*-score=1.732; B-H *p*-value=2.239×10^−02^) were all amongst the pathways upregulated in severe COVID-19 compared to mild COVID-19. HIF1α Signalling was also identified by IPA following treatment correction (IPA *z*-score=2.255; B-H *p*-value=3.890×10^−03^).

Inhibition of Angiogenesis by TSP1 was found to be downregulated by IPA whilst correcting for immunomodulatory treatment (IPA *z*-score=-0.258; B-H *p*-value=7.762×10^−03^), whilst PDGF signalling (*z*-score=1.414; B-H *p*-value=4.365×10^−02^) was found to be upregulated in severe COVID-19. Interestingly, the Th2 pathway (*z*-score=1.372; B-H *p*-value=1.549×10^−04^) was upregulated in severe COVID-19 whilst the Th1 pathway (*z*-score=-0.949; B-H *p*-value=7.413×10^−04^) was downregulated.

Following immunomodulatory treatment correction, there were several pathways downregulated in severe COVID-19 that were related to DNA damage and apoptosis, including NER (Nucleotide Excision Repair, Enhanced Pathway, *z*-score=-2.744; B-H *p*-value=1.318×10^−02^), Role of BRCA1 in DNA Damage Response (*z*-score=-0.943; B-H *p*-value=4.571×10^−02^) and TWEAK signalling (*z*-score=-0.5; B-H *p*-value=3.715×10^−02^). Furthermore, the translation pathways EIF2 Signalling (IPA *z*-score=-6.14; B-H *p*-value=1.259×10^−19^) and Regulation of eIF4 and p70S6K Signalling (IPA *z*-score=-0.728; B-H *p*-value=1.230×10^−08^) identified in the moderate *vs*. mild comparison remained significantly downregulated.

One pathway was enriched following immune cell proportion correction: Airway Pathology in Chronic Obstructive Pulmonary Disease (B-H *p*-value=4.571×10^−04^) with an indeterminate z-score.

#### Severe COVID-19 vs. Moderate COVID-19

8,971 genes were SDE between severe and moderate COVID-19 whilst accounting for immunomodulatory treatment with 4,380 and 4,591 genes over- and under-expressed with increasing severity, respectively. One gene (*NGFR*) was SDE between severe and moderate cases whilst accounting for immune cell proportions. *NGRF* was under-expressed in severe COVID-19 (B-H *p*-value=3.172×10^−02^; LFC=-1.692).

Inflammatory pathways were observed as upregulated by IPA (Supplementary Table 3), such as Natural Killer Cell Signalling (*z*-score=2.492; B-H *p*-value=1.000×10^−10^), IL-8 Signalling (*z*-score=4.341; B-H *p*-value=5.495×10^−05^), Acute Phase Response Signalling (*z*-score=4.523; B-H *p*-value=4.169×10^−04^) and IL-6 Signalling (*z*-score=4.32; B-H *p*-value=9.120×10^−06^).

Furthermore, multiple pathways related to macrophages were upregulated in severe COVID-19 compared to moderate COVID-19. These included production of Nitric Oxide and Reactive Oxygen Species in Macrophages (*z*-score= 3.571; B-H *p*-value=1.689×10^−04^), Fcγ Receptor-mediated Phagocytosis in Macrophages and Monocytes (*z*-score=2.959; B-H *p*-value=4.467×10^−05^) and Leukocyte Extravasation Signalling (*z*-score=2.219; B-H *p*-value=8.710×10^−06^).

Other pathways identified as upregulated in severe COVID-19 included Cardiac Hypertrophy Signalling (*z*-score=3.053; B-H *p*-value=1.318×10^−09^), Osteoarthritis Pathway (*z*-score=2.546; B-H *p*-value=4.074×10^−06^) and Neuroinflammation Signalling Pathway (*z*-score=2.557; B-H *p*-value=8.318×10^−07^). As observed in the severe *vs*. mild comparison, hypoxia related pathways were identified as enriched in severe COVID-19 including Hypoxia Signalling in the Cardiovascular System (*z*-score=1.732; B-H *p*-value=4.571×10^−02^) and HIF1α Signalling (*z*-score=2.109; B-H *p*-value=1.202×10^−07^).

Amongst the pathways downregulated in severe COVID-19 were various T cell pathways, including Th1 pathway (*z*-score=-0.302; B-H *p*-value=1.000×10^−10^), T Cell Receptor Signalling (*z*-score-7.553; B-H *p*-value=1.000×10^−06^), Systemic Lupus Erythematosus in T Cell Signalling Pathway (*z*-score:-2.286; B-H *p*-value=4.467×10^−05^) and Calcium-induced T Lymphocyte Apoptosis (*z*-score:-3.545; B-H *p*-value=2.951×10^−04^). PPAR signalling (*z*-score=-1.0915; B-H *p*-value=4.571×10^−04^) and PPARα/RXRα activation (*z*-score=-0.302; B-H *p*-value=3.548×10^−04^) were both downregulated in severe COVID-19, in addition to Antioxidant action of Vitamin C (*z*-score=-2.828; B-H *p*-value=3.548×10^−02^) and FAT10 Cancer Signalling Pathway (*z*-score=-1.807; B-H *p*-value=1.047×10^−02^).

#### COVID-19 severity as an additive variable

The impacts of severity on the transcriptome were also explored with severity as an additive variable (supplementary materials). 7,413 genes were SDE with severity whilst accounting for immunomodulatory treatment, of which 55 had absolute LFC values greater than 2 and adjusted *p--*values < 0.0001 (Fig. 3A). The 55 genes are split into two broad clusters (Fig. 3A) with the 1^st^ cluster containing multiple neutrophil-associated genes (including *LTF, MPO, BPI, ELANE*) and the 2^nd^ cluster, with two sub-clusters, containing various immunoglobulin and B cell genes (e.g., *IGHV3-13, IGKV6-1, IGHV3-10)*. 307 genes were SDE in all three pairwise severity comparisons whilst controlling for immunomodulatory treatment in addition to displaying additive behaviour (Supplementary Table 4). Of these 307, 10 genes had absolute LFC values greater than 2 and adjusted *p-*values < 0.0001 for the additive analyses (Fig. 3B). Genes SDE in the additive model in addition to the pairwise comparisons show more granularity than those SDE just in the additive model as their levels differ between each severity category in a stepwise manner (Supplementary Fig. 4).

**Figure 3.**
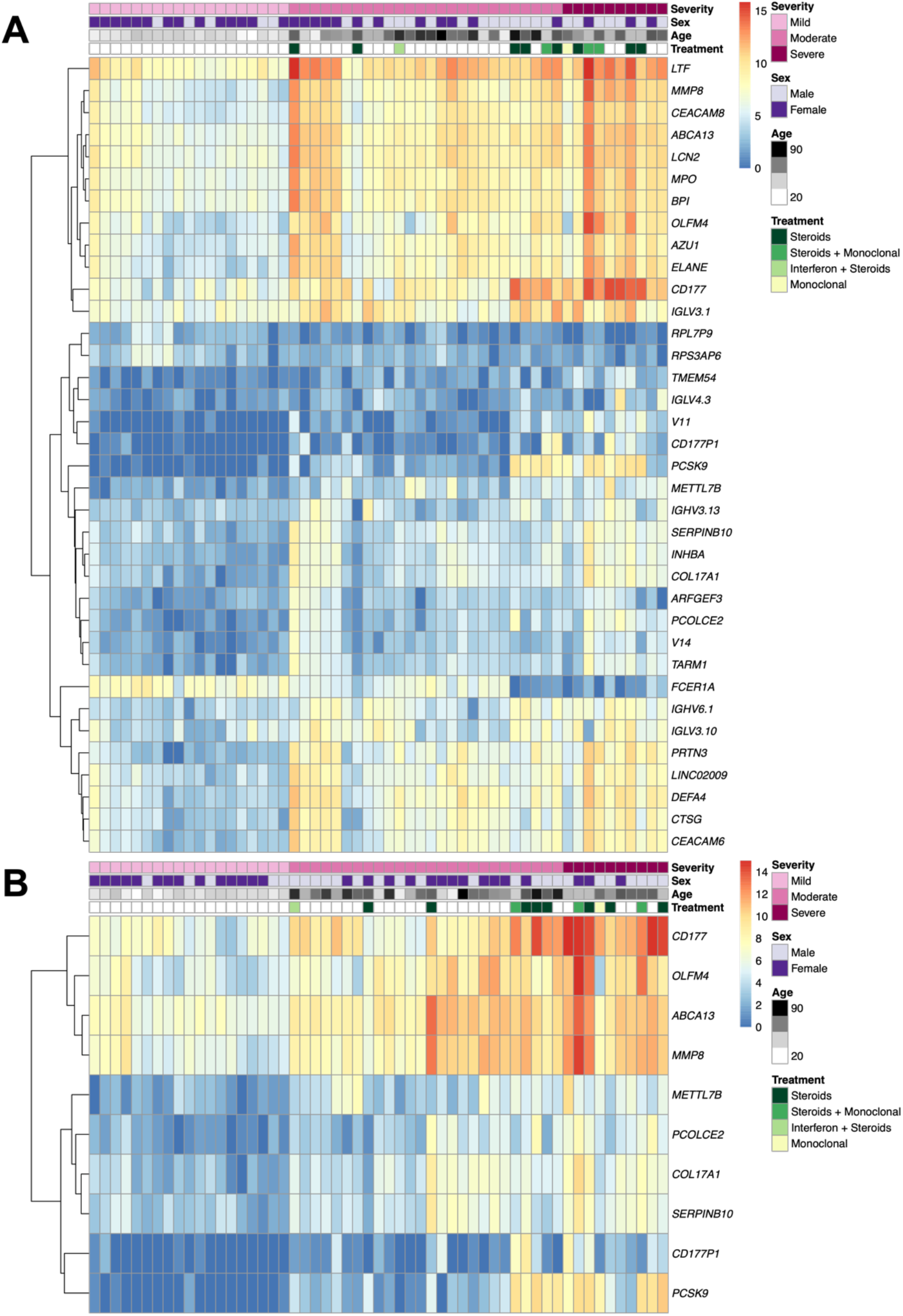
Heatmaps showing log-transformed expression values for **A:** the 55 genes SDE with severity as an additive variable with absolute LFC values greater than 2 and B-H *p*-values < 0.0001 **B:** the 10 genes SDE in all three pairwise severity comparisons in addition to the additive severity model with absolute LFC values greater than 2 and B-H *p*-values < 0.0001. Samples are ordered according to severity group.

## 5. Discussion

We have explored host whole-blood transcriptomes from COVID-19 patients with varying degrees of severity through differential expression and pathway analysis. We made pairwise comparisons between three different severity groups: mild, moderate, and severe. Severity analyses revealed major upregulation of genes and pathways related to the inflammatory immune response with increasing severity, with notable increases in genes and pathways related to neutrophil- and macrophage-mediated immunity accompanied by decreases in pathways related to T cell-mediated immunity.

We observed considerable transcriptomic differences between moderate COVID-19 individuals who received steroids to those who did not receive steroids, suggesting that this immunomodulatory treatment has a profound impact on the transcriptome. The widespread use of steroids in COVID-19 and the transcriptional disruption we observed in patients receiving steroids support the inclusion of immunomodulatory treatment in models related to COVID-19 transcriptomic analyses, in order to account for their impacts on the transcriptome.

Immune dysregulation has been extensively discussed as a contributing factor in the progression to severe COVID-19 ^5,8,36-38^. Infection with SARS-CoV-2 has been shown to induce a lower interferon response and an enhanced pro-inflammatory cytokine response in comparison to other viruses ^39^. This pro-inflammatory cytokine response leads to the attraction of monocytes and neutrophils, the development of a cytokine storm and hyperinflammation, and is likely to contribute to COVID-19 severity ^36,39^. Elevated levels of inflammatory cytokines and chemokines have been identified in plasma and serum from patients with increasing COVID-19 severity ^37,40,41^. We observed a domination of immune system associated pathways upregulated with increasing severity, with inflammatory immune pathways consistently identified.

Comparison of the *in-silico* estimates of immune cell proportions between the three severity groups revealed significant differences in all immune cell types except B cells. Lymphopenia was identified early in the pandemic as a key feature of COVID-19 severity ^42,43^. Our results reflect this as the levels of CD4 and CD8 T cells and NK cells reduce with increasing severity, whilst pathway analysis contrasting severe COVID-19 to either moderate or mild COVID-19 also revealed downregulation of many T cell related pathways.

Aschenbrenner et al. ^18^ observed neutrophil-specific gene expression changes with increasing COVID-19 severity in whole blood. We found that the proportions of *in silico* neutrophil estimations increased with severity. Furthermore, we identified various pathways related to neutrophils that increased with severity, such as TREM1 signalling, Inhibition of Matrix Metalloproteases and the STAT3 pathway. TREM1 has been associated with neutrophil migration across airway epithelial cells and has been suggested to increase inflammation through neutrophil migration into the lung ^44^. Matrix Metalloproteases (MMPs) are activated by neutrophil elastase and increased levels of multiple MMPs have been associated with increased COVID-19 severity ^45,46^.

To add to observations of upregulated neutrophil-related pathways and increased *in silico* neutrophil estimations with severity, Fig. 3A shows a clear cluster of neutrophil-associated genes with high absolute LFC values that were SDE with additive severity. When the blood transcriptomic profiles of mild COVID-19 were compared with moderate or severe COVID-19, *CEACAM8, MMP8, ELANE, LTF, CEACAM6* and *MPO* were consistently amongst the top SDE genes, with levels increasing with severity and high LFC. *MMP8* was also found to have an additive effect with severity. *ELANE, MPO* and *PRTN3*, which are linked to neutrophil degranulation and NETosis, have been found to be significantly altered in naso-oropharyngeal samples of SARS-CoV-2 infected patients ^47^. *ELANE, LTF, CEACAM8* and *MMP8* have been identified as being expressed in developing neutrophils, a novel cell subtype that was discovered through single-cell RNA sequencing of hospitalised COVID-19 patients, specifically identified in patients with acute respiratory distress syndrome (ARDS) ^48,49^. *CEACAM6* has been identified as having high expression in Type II pneumocytes in COVID-19 patients, the cells targeted by SARS-CoV-2 ^50^, and it has been suggested that cross-talk between Type II pneumocytes and developing neutrophils in COVID-19 occurs *via CEACAM8*-*CEACAM6* ^49^. It is possible that this cross-talk may promote differentiation of developing neutrophils, leading to further COVID-19 progression ^49^.

Pathways related to macrophage activation were identified with increasing COVID-19 severity. Increased levels of macrophage inflammatory protein 1α and monocyte-derived FCN1+ macrophage cells have been detected in individuals with more severe COVID-19 ^51,52^. The observations of increased inflammatory immune pathways and increased neutrophil and macrophage activity were accompanied with downregulation of T cell related pathways with increasing severity, including upregulation and downregulation of Th2 and Th1 pathways, respectively, with increasing severity. These findings suggest that the balance between different immune cell types is a key component that influences severity of COVID-19.

When immune cell proportions were included in the pairwise severity models, there was a considerable reduction in the number of SDE genes identified (Fig. 2), with only one gene identified as SDE between severe and moderate COVID-19. This observation indicates that much of the transcriptomic differences between individuals with varying severity of COVID-19 are driven by different proportions of immune cells. Furthermore, a large proportion of the pathways identified as enriched with severity were related to functions of different immune cells, suggesting that they play a major role in the pathogenesis of severe COVID-19.

Other pathways of interest include pathways related to hypoxia that were enriched with increasing severity. Hypoxia is a primary feature and major cause of mortality amongst patients with severe COVID-19 ^53^. Various pathways related to protein production and DNA/protein damage were identified as downregulated with increasing COVID-19 severity. SARS-CoV-2 has been shown to cause major disruption to host protein production, for example viral protein NSP1 has been shown to bind to the host 40S ribosomal subunit resulting in mRNA translation shut down ^54,55^. Furthermore, coronaviruses have also been shown to use DNA damage to induce cell cycle arrest ^56^. Damaged DNA can lead to accumulation of nuclear DNA in the cytoplasm which triggers innate immune responses ^57^.

Neutrophilia and neutrophil activation are usually markers of bacterial infection and are uncommon in most uncomplicated viral infections. In view of the association of increasing severity of COVID-19 with increased neutrophil counts, and expression of neutrophil genes including those involved in degranulation, NETosis, and neutrophil-mediated tissue injury, a key question is what mechanisms are responsible for the shift from the normal “viral” response in mild COVID-19, to the severe inflammatory process involving neutrophils in severe disease. The timing of the inflammatory phase of COVID-19 usually occurring in the second week of illness ^58,59^, together with increased expression of immunoglobulin genes we observed (e.g., *IGKV1D-13, IGHV3-43, IGLV4-3, IGLV3-16, IGLV3-10*) may suggest that immunoglobulin, directed at either viral or modified self-antigens, may be involved in neutrophil and macrophage activation through Fc-gamma receptors or complement mediated activation, genes related to which are upregulated in severe disease (e.g., *FCGR2A, FCGR3B, FCGR3A, CR1, C3AR1, C5AR1*). Neutrophil activation and neutrophil-mediated tissue injury may be a promising target for therapeutic intervention.

### 5.1. Limitations

COVID-19 severity is highly influenced by age which leads to major confounding between these two variables. Although we controlled for this confounder (by including age and the interaction between age and severity) when exploring transcriptomic changes with severity, it is possible that we may have a) failed to identify key drivers of severity as they are confounded with age, b) inadvertently included spurious genes that are really driven by age rather than severity. The sample sizes in our analyses are modest for some severity groups. For example, in the severe COVID-19 group, only 10 samples could be included due to concomitant bacterial infection, because coinfections were likely to have had profound transcriptional impacts and may have masked the genuine SARS-CoV-2 signal.

### 5.2. Conclusion

We have explored the transcriptomic impact of SARS-CoV-2 infection through evaluating the transcriptomic differences between individuals with varying levels of COVID-19 severity. We have observed considerable transcriptomic perturbation which offer insights into the host factors that influence development of severe COVID-19. Upregulation of inflammatory immune pathways was observed with increasing severity, with multiple neutrophil, macrophage and immunoglobulin-associated genes and pathways identified, suggesting that increased COVID-19 severity may be mediated in part by neutrophil activation, which may be related to production of immunoglobulin as acquired immunity develops.

## Data Availability

Gene counts and sample metadata are available on ArrayExpress under the accession E-MTAB-10926.

## Author Contributions

Conceptualization: MK, FM-T, ML; Data curation: IRC, HJ, VJW, AG-C, DH-C; Formal analysis: HJ, DH-C; funding acquisition: HJ, CB, VJW, JAH, ML, AJC, MK, FM-T, IRC, AG-C, AS; Investigation: IRC, AG-C, VJW, SN, GS, DH-C; Methodology: HJ, VJW, DH-C; Project administration: IRC, MK, VJW, AS, AG-C, FM-T; Resources: FM-T, IRC, VJW, AG-C, GS, SN, MK; Software: HJ, DH-C; Supervision: MK, VJW, JAH, ML, FM-T; Validation: CB; Visualisation: HJ; Writing—original draft: HJ, IRC, MK; Writing—review and editing: ALL.

**All authors have read and agreed to the published version of the manuscript**.

## Acknowledgments

Many thanks to the patients and their families who took part in the study that the data presented here originated from.

