## Supplementary materials for "Characterisation of the blood RNA host response underpinning severity in COVID-19 patients"

1. ***Supplementary Methods***

Severity was explored as a numerical variable in an additive model with 0, 1 and 2 values indicating mild, moderate, and severe cases, respectively, to identify genes that are significantly differentially expressed (SDE) across the three severity groups sequentially. The analysis was repeated twice with different model designs. The first model design aimed to account for transcriptomic differences induced by immunomodulatory treatments. This model included parameters representing whether the patients received tocilizumab, steroids or interferon treatments, in addition to sex, age, severity. The second model design accounted for transcriptomic differences induced by different proportions of immune cells. The immune cell proportions accounted for included: monocytes, neutrophils, B cells (the sum of naïve and memory B cells and plasma cell proportions), CD4 T cells, (the sum of the proportions of naïve CD4 T cells, resting and activated memory CD4 T cells, follicular helper T cells and regulatory T cells), CD8 T cells and natural killer cells (the sum of resting and activated natural killer [NK] cell proportions). In addition to the immune cell proportions, this model included sex, age, severity. Pathway analysis using Ingenuity Pathway Analysis (IPA; QIAGEN Inc., <https://www.qiagenbioinformatics.com/products/ingenuity-pathway-analysis>) was performed on the SDE genes. Genes SDE with age were also identified through using a DESeq2 model with age, severity (as an additive variable) the interaction between and severity and sex. This model designed to identify genes SDE with age that could be excluded due to confounding between age and severity.

2. ***Supplementary Results***
   2.1. *The effect of immunomodulatory treatment on COVID-19 patients’ blood transcriptome*

The impacts of steroid administration on the transcriptome were explored through contrasting moderate COVID-19 patients who received steroids (*n*=6) to moderate COVID-19 patients who did not receive steroids (*n*=20). IPA identified no significant pathways from the SDE genes. Full results in File_S1_Steroids.xlsx.

2.2. *Transcriptomic differences between different COVID-19 severity groups*

*Moderate COVID-19 vs*. *Mild COVID-19*

Full lists of SDE genes and pathways are in Supplementary File 2 (File_S2_Moderate_vs_Mild_COVID19.xlsx). IPA identified 24 significant pathways (Supplementary Table 1) from the list of genes SDE between moderate and mild COVID-19 whilst accounting for immunomodulatory treatment (*n*=1,547), with 9 and 12 pathways up and downregulated, respectively. IPA identified two significant pathways (EIF2 Signalling: *z*-score=-2.53, B-H *p*-value=1.288×10^-03^; Regulation of eIF4 and p70S6K Signalling: B-H *p*-value=1.950×10^-02^) from the list of genes SDE between moderate and mild COVID-19 whilst accounting for immune cell proportions (*n*=488).

*Severe COVID-19 vs. Mild COVID-19*

Full lists of SDE genes and pathways are in Supplementary File 3 (File_S3_Severe_vs_Mild_COVID19.xlsx). IPA identified 68 significant pathways (Supplementary Table 2) from the list of genes SDE between severe COVID-19 and mild COVID-19 whilst accounting for immunomodulatory treatment (*n*=7,343), with 33 and 19 pathways upregulated and downregulated, respectively. When immune cell proportions were included in the model instead of immunomodulatory treatment, IPA identified one significant pathway (Airway Pathology in Chronic Obstructive Pulmonary Disease, B-H *p*-value=4.571×10^-04^) from the list of genes SDE (*n*=94).

*Severe COVID-19 vs*. *Moderate COVID-19*

Full lists of SDE genes and pathways are in Supplementary File 4 (File_S4_Severe_vs_Moderate_COVID19.xlsx). IPA identified 260 significant pathways (Supplementary Table 3) from the list of genes SDE between severe COVID-19 and moderate COVID-19 whilst accounting for immunomodulatory treatment (*n*=8,971) with 179 and 23 pathways up and down regulated, respectively. When immune cell proportions were included in the model instead of immunomodulatory treatment, one SDE genes was identified (*NGRF*) so pathway analysis was not performed.

2.3. *Additive Severity Model*

In addition to exploring the transcriptomic differences between the pairwise severity groups, we also explored severity as an additive variable to attempt to identify genes that are SDE across all three groups sequentially. The full lists of genes and pathways are in Supplementary File 5 (File_S5_Additive_Severity.xlsx). The models exploring severity as an additive variable were ran twice; once with immunomodulatory treatment variables included in the model, and once with immune cell proportions included in the model. When the immunomodulatory treatments were accounted for the in DESeq2 model, there were 7,414 genes SDE (adjusted *p*-value < 0.05) with severity. 3,626 genes were over-expressed with severity and 3,788 genes were under-expressed with increasing severity. When the cell proportions were accounted for the in DESeq2 model, there were 88 genes SDE (adjusted *p*-value < 0.05) with severity. 74 genes were over-expressed with severity and 14 genes were under-expressed with increasing severity. Whilst 82 genes were SDE in both comparisons (Supplementary Fig. 3), the treatment and immune cell corrections revealed 7,330 and 6 additional SDE genes, respectively.

IPA pathway analyses were performed. When immunomodulatory treatment was included in the model, 123 significant pathways were identified by IPA with 84 and 19 pathways increasing and decreasing with severity, respectively (Supplementary Table 4). When cell proportions were included in the model, one pathway was identified by IPA: Airway Pathology in Chronic Obstructive Pulmonary Disease (B-H *p*-value=3.89×10^-04^).

307 genes were identified as having additive behaviour in addition to being SDE in pairwise analyses whilst correcting for immunomodulatory treatment (Supplementary Table 5). Of the 307 genes, all log_2_ fold-change directions were concordant with 96 and 211 genes increasing and decreasing with severity, respectively.

2.4. *Identification of genes SDE with age*

We used DESeq2 to identify genes SDE with age in COVID-19 patients in our dataset. The model included sex, age, severity, and the interaction between age and severity. We did this to see whether there would be genes that could be SDE with severity but were not identified due to their associations with age. 25 genes were SDE with age (Supplementary Table 6).

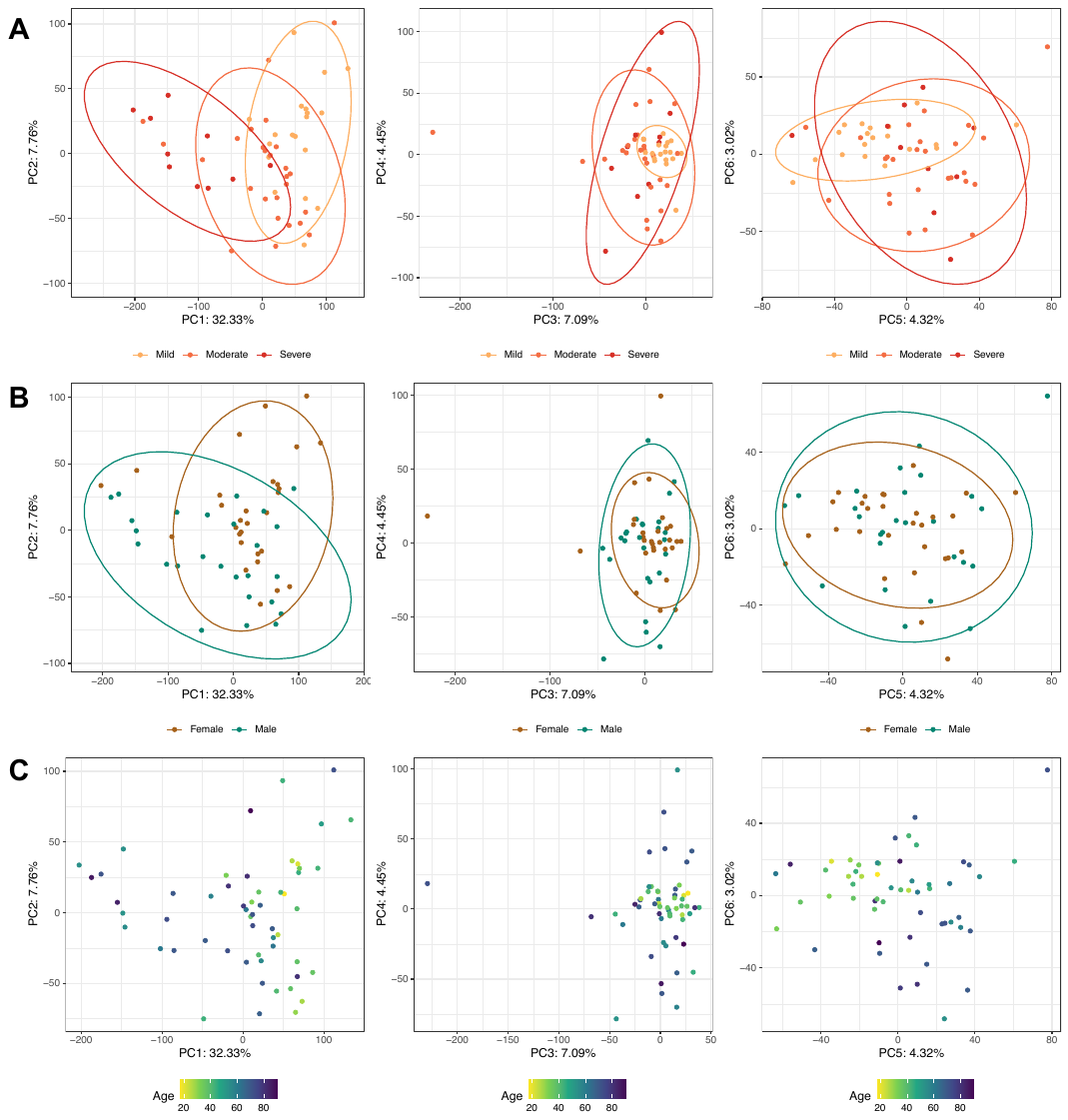
Supplementary Figure 1 Principal component analysis (PCA) plots for the samples used in the analyses. Points, which represent samples, are coloured by disease severity (A), sex (B) and age (C).

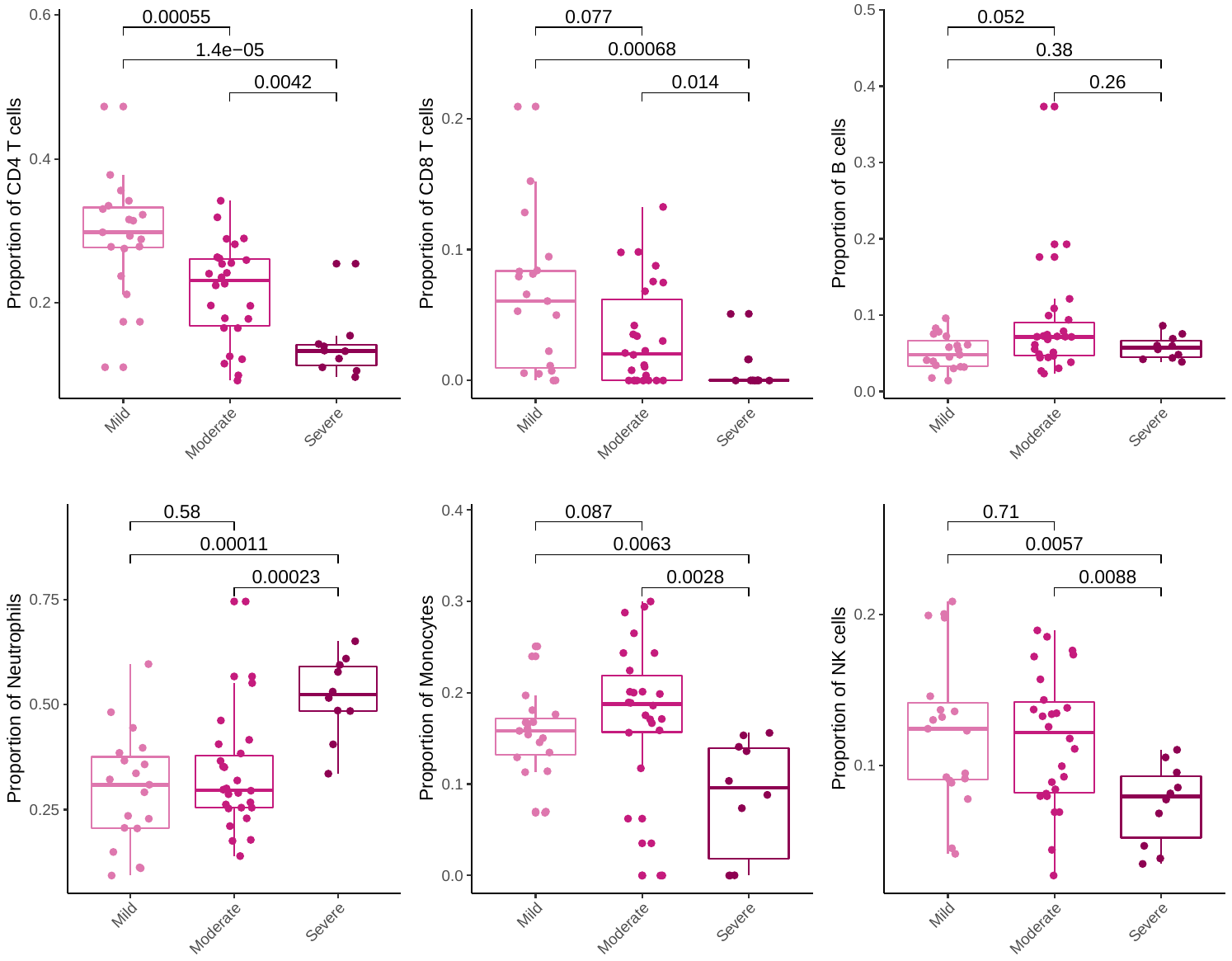

Supplementary Figure 2 *In silico* predicted immune cell proportions for COVID-19 samples across severity groups. Immune cell proportions were estimated using CIBERSORTx [1]. B cells are the sum of naïve and memory B cells and plasma cell proportions, CD4 T cells are the sum of naïve CD4 T cells, resting and activated memory CD4 T cells, follicular helper T cells and regulatory T cells proportions, and natural killer cells are the sum of resting and activated NK cell proportions. P-values are calculated using two-sided Mann-Whitney-Wilcoxon with Benjamini-Hochberg correction.

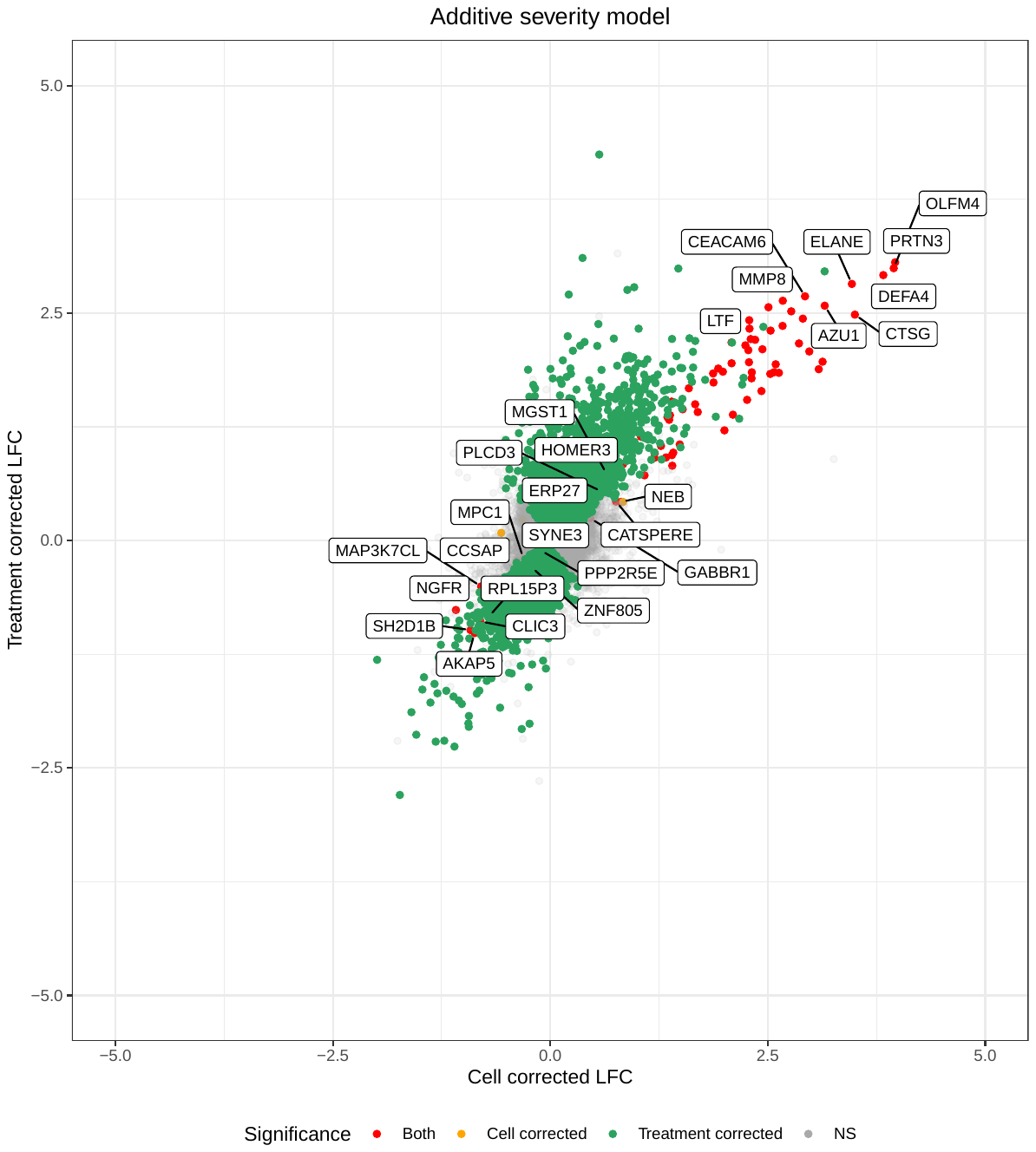

**Supplementary Figure 3** Cross plot showing the log2 fold change (LFC) values of genes with severity as an additive variable and how LFC values differ according to whether immune cell proportions (x-axis) or immunomodulatory treatments (y-axis) are included in the model. Red points are genes that are were SDE in both models, whilst orange and green points are genes SDE in the cell correction and treatment correction models, respectively. NS = not significant.

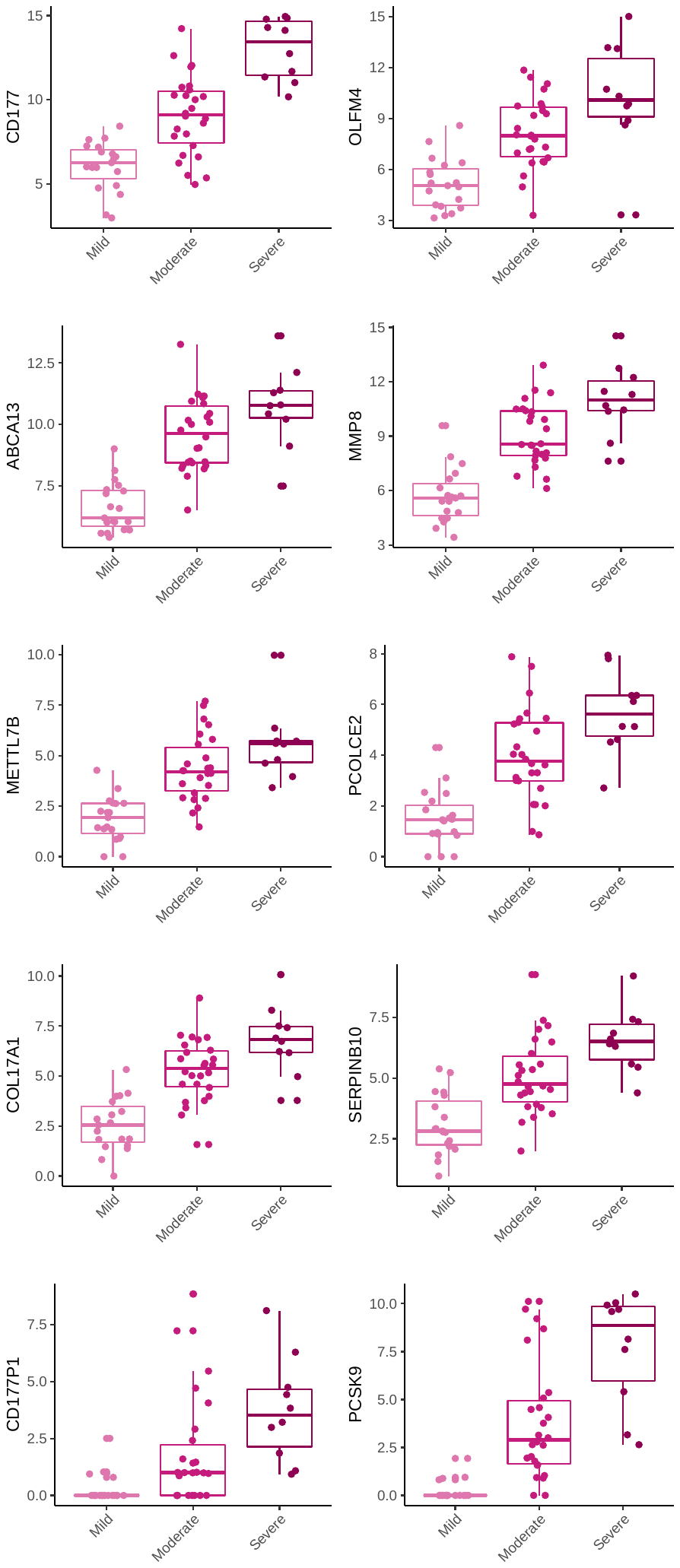

**Supplementary Figure 4** Boxplots for the top genes SDE in the additive severity analysis in addition to each pairwise comparison from the DESeq2 models including age, sex and immunomodulatory treatment. These genes had absolute LFC values greater than 2 and adjusted p-values < 0.0001 in the additive DESeq2 model and are also shown in Fig. 3B. Counts are logged and normalised.

**Supplementary Table 1** The top 15 pathways (from a total of 24 significant pathways) identified by IPA from the genes SDE between moderate COVID-19 *vs*. mild COVID-19 (B-H p-value < 0.05) whilst correcting for immunomodulatory treatment. Positive and negative Z-scores indicate pathway upregulation and downregulation, respectively, in moderate COVID-19 *vs*. mild COVID-19.

| **Ingenuity Canonical Pathways** | **B-H p-value** | **Z-score** |
| --- | --- | --- |
| EIF2 Signaling | 5.012×10^-29^ | -5.778 |
| Regulation of eIF4 and p70S6K Signaling | 1.000×10^-13^ | -1 |
| mTOR Signaling | 4.467×10^-08^ | -0.632 |
| Coronavirus Pathogenesis Pathway | 5.248×10^-07^ | 2.744 |
| Kinetochore Metaphase Signaling Pathway | 1.820×10^-05^ | 0 |
| Mitotic Roles of Polo-Like Kinase | 2.754×10^-05^ | 0.816 |
| Oxidative Phosphorylation | 1.514×10^-03^ | -4.243 |
| Unfolded protein response | 1.514×10^-03^ | -1 |
| Mitochondrial Dysfunction | 2.692×10^-03^ | NA |
| Cell Cycle Regulation by BTG Family Proteins | 5.888×10^-03^ | NA |
| Sirtuin Signaling Pathway | 5.888×10^-03^ | 0.218 |
| Huntington's Disease Signaling | 6.026×10^-03^ | 0.302 |
| Role of CHK Proteins in Cell Cycle Checkpoint Control | 7.943×10^-03^ | -2.828 |
| ATM Signaling | 7.943×10^-03^ | 0 |
| Spliceosomal Cycle | 8.318×10^-03^ | -3.162 |

**Supplementary Table 2** The top 15 (from a total of 68 significant pathways) identified by IPA from the genes SDE between severe COVID-19 *vs*. mild COVID-19 (B-H p-value < 0.05) whilst correcting for immunomodulatory treatment. Positive and negative Z-scores indicate pathway upregulation and downregulation, respectively, in severe COVID-19 *vs*. mild COVID-19.

| **Ingenuity Canonical Pathways** | **B-H p-value** | **Z-score** |
| --- | --- | --- |
| EIF2 Signaling | 1.259×10^-19^ | -6.14 |
| Regulation of eIF4 and p70S6K Signaling | 1.230×10^-08^ | -0.728 |
| mTOR Signaling | 9.333×10^-06^ | -1 |
| Coronavirus Pathogenesis Pathway | 9.333×10^-06^ | 3.667 |
| Th1 and Th2 Activation Pathway | 1.820×10^-05^ | NA |
| Th2 Pathway | 1.549×10^-04^ | 1.372 |
| Th1 Pathway | 7.413×10^-04^ | -0.949 |
| Natural Killer Cell Signaling | 7.413×10^-04^ | 0 |
| 4-1BB Signaling in T Lymphocytes | 9.120×10^-04^ | 0.728 |
| Role of PKR in Interferon Induction and Antiviral Response | 9.120×10^-04^ | 1.54 |
| B Cell Receptor Signaling | 1.479×10^-03^ | 1.408 |
| RANK Signaling in Osteoclasts | 2.754×10^-03^ | 1.715 |
| Role of CHK Proteins in Cell Cycle Checkpoint Control | 3.890×10^-03^ | -2.236 |
| Unfolded protein response | 3.890×10^-03^ | 1.043 |
| HIF1α Signaling | 3.890×10^-03^ | 2.255 |

**Supplementary Table 3** The top 15 pathways (from a total of 260 significant pathways) by IPA from the genes SDE between severe COVID-19 *vs*. moderate COVID-19 (B-H p-value < 0.05) whilst correcting for immunomodulatory treatment. Positive and negative Z-scores indicate pathway upregulation and downregulation, respectively, in severe COVID-19 *vs*. moderate COVID-19.

| **Ingenuity Canonical Pathways** | **B-H p-value** | **Z-score** |
| --- | --- | --- |
| Th1 and Th2 Activation Pathway | 1.995×10^-12^ | NA |
| Th1 Pathway | 1.000×10^-10^ | -0.302 |
| Natural Killer Cell Signaling | 1.000×10^-10^ | 2.492 |
| Th2 Pathway | 2.291×10^-10^ | 1.441 |
| STAT3 Pathway | 3.388×10^-10^ | 1.808 |
| TREM1 Signaling | 6.310×10^-10^ | 4.218 |
| Cardiac Hypertrophy Signaling (Enhanced) | 1.318×10^-09^ | 3.053 |
| Hepatic Fibrosis Signaling Pathway | 1.023×10^-08^ | 2.376 |
| Role of Macrophages, Fibroblasts and Endothelial Cells in Rheumatoid Arthritis | 3.631×10^-08^ | NA |
| NF-κB Signaling | 1.072×10^-07^ | 2.066 |
| HIF1α Signaling | 1.202×10^-07^ | 2.109 |
| Tec Kinase Signaling | 1.202×10^-07^ | 2.111 |
| Axonal Guidance Signaling | 1.202×10^-07^ | NA |
| PI3K/AKT Signaling | 2.512×10^-07^ | 1.095 |
| HGF Signaling | 2.512×10^-07^ | 2.744 |

**Supplementary Table 4** The top 15 pathways (from a total of 123 significant pathways) by IPA from the genes SDE with severity as an additive variable whilst correcting for immunomodulatory treatment. Positive and negative Z-scores indicate pathway upregulation and downregulation, respectively, with increasing COVID-19 severity.

| **Ingenuity Canonical Pathways** | **B-H p-value** | **Z-score** |
| --- | --- | --- |
| EIF2 Signaling | 2.512×10^-22^ | -5.421 |
| Regulation of eIF4 and p70S6K Signaling | 2.512×10^-11^ | 0.447 |
| Coronavirus Pathogenesis Pathway | 2.455×10^-07^ | 4.013 |
| mTOR Signaling | 9.120×10^-07^ | 0 |
| B Cell Receptor Signaling | 9.120×10^-07^ | 2.492 |
| Th1 and Th2 Activation Pathway | 1.479×10^-06^ | NA |
| Natural Killer Cell Signaling | 2.630×10^-06^ | 0.671 |
| Th1 Pathway | 5.623×10^-05^ | -0.152 |
| Th2 Pathway | 5.623×10^-05^ | 1.809 |
| STAT3 Pathway | 3.802×10^-04^ | 1.508 |
| HGF Signaling | 8.128×10^-04^ | 1.581 |
| Role of PKR in Interferon Induction and Antiviral Response | 9.120×10^-04^ | 2.1 |
| HIF1α Signaling | 9.772×10^-04^ | 2.79 |
| PPAR Signaling | 1.413×10^-03^ | -0.457 |
| RANK Signaling in Osteoclasts | 2.630×10^-03^ | 2.058 |

**Supplementary Table 5** Genes with additive behaviour in addition to being significant in all pairwise comparisons whilst correcting for immunomodulatory treatment. Direction of ‘up’ means their levels increase with increasing severity.

| **ID** | **Gene** | **Direction** |
| --- | --- | --- |
| ENSG00000204933 | CD177P1 | Up |
| ENSG00000169174 | PCSK9 | Up |
| ENSG00000102837 | OLFM4 | Up |
| ENSG00000204936 | CD177 | Up |
| ENSG00000118113 | MMP8 | Up |
| ENSG00000163710 | PCOLCE2 | Up |
| ENSG00000179869 | ABCA13 | Up |
| ENSG00000065618 | COL17A1 | Up |
| ENSG00000170439 | METTL7B | Up |
| ENSG00000242550 | SERPINB10 | Up |
| ENSG00000064270 | ATP2C2 | Up |
| ENSG00000268833 | - | Up |
| ENSG00000211965 | IGHV3-49 | Up |
| ENSG00000069535 | MAOB | Up |
| ENSG00000211665 | IGLV3-16 | Up |
| ENSG00000211937 | IGHV2-5 | Up |
| ENSG00000160180 | TFF3 | Up |
| ENSG00000224940 | PRRT4 | Up |
| ENSG00000241244 | IGKV1D-16 | Up |
| ENSG00000232216 | IGHV3-43 | Up |
| ENSG00000136010 | ALDH1L2 | Up |
| ENSG00000211959 | IGHV4-39 | Up |
| ENSG00000139572 | GPR84 | Up |
| ENSG00000278196 | IGLV2-8 | Up |
| ENSG00000224650 | IGHV3-74 | Up |
| ENSG00000104918 | RETN | Up |
| ENSG00000279400 | - | Up |
| ENSG00000182175 | RGMA | Up |
| ENSG00000106853 | PTGR1 | Up |
| ENSG00000133063 | CHIT1 | Up |
| ENSG00000158352 | SHROOM4 | Up |
| ENSG00000005961 | ITGA2B | Up |
| ENSG00000079393 | DUSP13 | Up |
| ENSG00000275898 | - | Up |
| ENSG00000164850 | GPER1 | Up |
| ENSG00000250280 | - | Up |
| ENSG00000163661 | PTX3 | Up |
| ENSG00000265688 | MAFG-DT | Up |
| ENSG00000168528 | SERINC2 | Up |
| ENSG00000122861 | PLAU | Up |
| ENSG00000162407 | PLPP3 | Up |
| ENSG00000070371 | CLTCL1 | Up |
| ENSG00000109265 | CRACD | Up |
| ENSG00000137563 | GGH | Up |
| ENSG00000228923 | - | Up |
| ENSG00000197653 | DNAH10 | Up |
| ENSG00000250644 | - | Up |
| ENSG00000100024 | UPB1 | Up |
| ENSG00000135862 | LAMC1 | Up |
| ENSG00000223916 | - | Up |
| ENSG00000105711 | SCN1B | Up |
| ENSG00000111186 | WNT5B | Up |
| ENSG00000144407 | PTH2R | Up |
| ENSG00000184702 | SEPTIN5 | Up |
| ENSG00000226928 | RPS14P4 | Up |
| ENSG00000279276 | - | Up |
| ENSG00000051128 | HOMER3 | Up |
| ENSG00000152380 | FAM151B | Up |
| ENSG00000185909 | KLHDC8B | Up |
| ENSG00000197063 | MAFG | Up |
| ENSG00000280167 | - | Up |
| ENSG00000008394 | MGST1 | Up |
| ENSG00000111181 | SLC6A12 | Up |
| ENSG00000072954 | TMEM38A | Up |
| ENSG00000104043 | ATP8B4 | Up |
| ENSG00000228817 | BACH1-IT2 | Up |
| ENSG00000229512 | - | Up |
| ENSG00000064601 | CTSA | Up |
| ENSG00000075651 | PLD1 | Up |
| ENSG00000258337 | - | Up |
| ENSG00000118985 | ELL2 | Up |
| ENSG00000271795 | - | Up |
| ENSG00000108405 | P2RX1 | Up |
| ENSG00000164236 | ANKRD33B | Up |
| ENSG00000221598 | MIR1249 | Up |
| ENSG00000248476 | BACH1-IT1 | Up |
| ENSG00000199805 | RNU1-134P | Up |
| ENSG00000263120 | - | Up |
| ENSG00000262580 | - | Up |
| ENSG00000277255 | MIR7854 | Up |
| ENSG00000227200 | - | Up |
| ENSG00000067225 | PKM | Up |
| ENSG00000127838 | PNKD | Up |
| ENSG00000279476 | - | Up |
| ENSG00000181523 | SGSH | Up |
| ENSG00000243508 | DNAJB6P7 | Up |
| ENSG00000198018 | ENTPD7 | Up |
| ENSG00000267632 | - | Up |
| ENSG00000070214 | SLC44A1 | Up |
| ENSG00000162341 | TPCN2 | Up |
| ENSG00000166340 | TPP1 | Up |
| ENSG00000198113 | TOR4A | Up |
| ENSG00000278949 | - | Up |
| ENSG00000165714 | BORCS5 | Up |
| ENSG00000196295 | GARS1-DT | Up |
| ENSG00000164961 | WASHC5 | Up |
| ENSG00000154001 | PPP2R5E | Down |
| ENSG00000253719 | ATXN7L3B | Down |
| ENSG00000183513 | COA5 | Down |
| ENSG00000172262 | ZNF131 | Down |
| ENSG00000165943 | MOAP1 | Down |
| ENSG00000139163 | ETNK1 | Down |
| ENSG00000149311 | ATM | Down |
| ENSG00000107771 | CCSER2 | Down |
| ENSG00000149313 | AASDHPPT | Down |
| ENSG00000198252 | STYX | Down |
| ENSG00000198791 | CNOT7 | Down |
| ENSG00000186104 | CYP2R1 | Down |
| ENSG00000047932 | GOPC | Down |
| ENSG00000173726 | TOMM20 | Down |
| ENSG00000104979 | C19orf53 | Down |
| ENSG00000119335 | SET | Down |
| ENSG00000066654 | THUMPD1 | Down |
| ENSG00000204977 | TRIM13 | Down |
| ENSG00000110696 | C11orf58 | Down |
| ENSG00000138767 | CNOT6L | Down |
| ENSG00000150593 | PDCD4 | Down |
| ENSG00000139154 | AEBP2 | Down |
| ENSG00000144034 | TPRKB | Down |
| ENSG00000122034 | GTF3A | Down |
| ENSG00000083535 | PIBF1 | Down |
| ENSG00000080822 | CLDND1 | Down |
| ENSG00000116750 | UCHL5 | Down |
| ENSG00000129317 | PUS7L | Down |
| ENSG00000163322 | ABRAXAS1 | Down |
| ENSG00000126804 | ZBTB1 | Down |
| ENSG00000115419 | GLS | Down |
| ENSG00000147679 | UTP23 | Down |
| ENSG00000148362 | PAXX | Down |
| ENSG00000107625 | DDX50 | Down |
| ENSG00000189227 | C15orf61 | Down |
| ENSG00000198331 | HYLS1 | Down |
| ENSG00000137770 | CTDSPL2 | Down |
| ENSG00000165156 | ZHX1 | Down |
| ENSG00000122484 | RPAP2 | Down |
| ENSG00000163607 | GTPBP8 | Down |
| ENSG00000171490 | RSL1D1 | Down |
| ENSG00000167842 | MIS12 | Down |
| ENSG00000170903 | MSANTD4 | Down |
| ENSG00000170364 | SETMAR | Down |
| ENSG00000214367 | HAUS3 | Down |
| ENSG00000197894 | ADH5 | Down |
| ENSG00000166037 | CEP57 | Down |
| ENSG00000161016 | RPL8 | Down |
| ENSG00000251022 | THAP9-AS1 | Down |
| ENSG00000122873 | CISD1 | Down |
| ENSG00000144895 | EIF2A | Down |
| ENSG00000106460 | TMEM106B | Down |
| ENSG00000204387 | SNHG32 | Down |
| ENSG00000162244 | RPL29 | Down |
| ENSG00000005469 | CROT | Down |
| ENSG00000255559 | ZNF252P-AS1 | Down |
| ENSG00000162623 | TYW3 | Down |
| ENSG00000117906 | RCN2 | Down |
| ENSG00000111875 | ASF1A | Down |
| ENSG00000188846 | RPL14 | Down |
| ENSG00000151332 | MBIP | Down |
| ENSG00000197056 | ZMYM1 | Down |
| ENSG00000105193 | RPS16 | Down |
| ENSG00000166226 | CCT2 | Down |
| ENSG00000182141 | ZNF708 | Down |
| ENSG00000226287 | TMEM191A | Down |
| ENSG00000273015 | - | Down |
| ENSG00000074935 | TUBE1 | Down |
| ENSG00000164022 | AIMP1 | Down |
| ENSG00000254838 | GVINP1 | Down |
| ENSG00000256087 | ZNF432 | Down |
| ENSG00000139324 | TMTC3 | Down |
| ENSG00000100442 | FKBP3 | Down |
| ENSG00000105829 | BET1 | Down |
| ENSG00000090612 | ZNF268 | Down |
| ENSG00000088179 | PTPN4 | Down |
| ENSG00000139343 | SNRPF | Down |
| ENSG00000133641 | C12orf29 | Down |
| ENSG00000138660 | AP1AR | Down |
| ENSG00000156017 | CARNMT1 | Down |
| ENSG00000213186 | TRIM59 | Down |
| ENSG00000109971 | HSPA8 | Down |
| ENSG00000180257 | ZNF816 | Down |
| ENSG00000180917 | CMTR2 | Down |
| ENSG00000120694 | HSPH1 | Down |
| ENSG00000245910 | SNHG6 | Down |
| ENSG00000110700 | RPS13 | Down |
| ENSG00000115816 | CEBPZ | Down |
| ENSG00000182287 | AP1S2 | Down |
| ENSG00000090266 | NDUFB2 | Down |
| ENSG00000174748 | RPL15 | Down |
| ENSG00000114686 | MRPL3 | Down |
| ENSG00000166275 | BORCS7 | Down |
| ENSG00000149273 | RPS3 | Down |
| ENSG00000083845 | RPS5 | Down |
| ENSG00000120686 | UFM1 | Down |
| ENSG00000197958 | RPL12 | Down |
| ENSG00000134049 | IER3IP1 | Down |
| ENSG00000170846 | - | Down |
| ENSG00000164172 | MOCS2 | Down |
| ENSG00000221944 | TIGD1 | Down |
| ENSG00000177721 | ANXA2R | Down |
| ENSG00000058729 | RIOK2 | Down |
| ENSG00000233757 | - | Down |
| ENSG00000182359 | KBTBD3 | Down |
| ENSG00000018869 | ZNF582 | Down |
| ENSG00000147403 | RPL10 | Down |
| ENSG00000105849 | POLR1F | Down |
| ENSG00000144713 | RPL32 | Down |
| ENSG00000158691 | ZSCAN12 | Down |
| ENSG00000075089 | ACTR6 | Down |
| ENSG00000167232 | ZNF91 | Down |
| ENSG00000198440 | ZNF583 | Down |
| ENSG00000180787 | ZFP3 | Down |
| ENSG00000152133 | GPATCH11 | Down |
| ENSG00000106591 | MRPL32 | Down |
| ENSG00000123545 | NDUFAF4 | Down |
| ENSG00000241127 | YAE1 | Down |
| ENSG00000163577 | EIF5A2 | Down |
| ENSG00000104408 | EIF3E | Down |
| ENSG00000113966 | ARL6 | Down |
| ENSG00000197050 | ZNF420 | Down |
| ENSG00000259834 | - | Down |
| ENSG00000168283 | BMI1 | Down |
| ENSG00000083099 | LYRM2 | Down |
| ENSG00000254004 | ZNF260 | Down |
| ENSG00000255135 | - | Down |
| ENSG00000198707 | CEP290 | Down |
| ENSG00000177888 | ZBTB41 | Down |
| ENSG00000168116 | KIAA1586 | Down |
| ENSG00000151835 | SACS | Down |
| ENSG00000137038 | DMAC1 | Down |
| ENSG00000172172 | MRPL13 | Down |
| ENSG00000198346 | ZNF813 | Down |
| ENSG00000168028 | RPSA | Down |
| ENSG00000143971 | ETAA1 | Down |
| ENSG00000169288 | MRPL1 | Down |
| ENSG00000174444 | RPL4 | Down |
| ENSG00000163281 | GNPDA2 | Down |
| ENSG00000179144 | GIMAP7 | Down |
| ENSG00000101132 | PFDN4 | Down |
| ENSG00000186468 | RPS23 | Down |
| ENSG00000142937 | RPS8 | Down |
| ENSG00000100316 | RPL3 | Down |
| ENSG00000137501 | SYTL2 | Down |
| ENSG00000120526 | NUDCD1 | Down |
| ENSG00000140006 | WDR89 | Down |
| ENSG00000116791 | CRYZ | Down |
| ENSG00000142541 | RPL13A | Down |
| ENSG00000198464 | ZNF480 | Down |
| ENSG00000177932 | ZNF354C | Down |
| ENSG00000182774 | RPS17 | Down |
| ENSG00000122406 | RPL5 | Down |
| ENSG00000136897 | MRPL50 | Down |
| ENSG00000196911 | KPNA5 | Down |
| ENSG00000261366 | MANEA-DT | Down |
| ENSG00000236552 | RPL13AP5 | Down |
| ENSG00000164587 | RPS14 | Down |
| ENSG00000152219 | ARL14EP | Down |
| ENSG00000197841 | ZNF181 | Down |
| ENSG00000118181 | RPS25 | Down |
| ENSG00000067840 | PDZD4 | Down |
| ENSG00000167286 | CD3D | Down |
| ENSG00000163519 | TRAT1 | Down |
| ENSG00000156508 | EEF1A1 | Down |
| ENSG00000111678 | C12orf57 | Down |
| ENSG00000174946 | GPR171 | Down |
| ENSG00000142875 | PRKACB | Down |
| ENSG00000078596 | ITM2A | Down |
| ENSG00000143947 | RPS27A | Down |
| ENSG00000169740 | ZNF32 | Down |
| ENSG00000008988 | RPS20 | Down |
| ENSG00000244720 | NT5C3AP2 | Down |
| ENSG00000146757 | ZNF92 | Down |
| ENSG00000164114 | MAP9 | Down |
| ENSG00000213741 | RPS29 | Down |
| ENSG00000137154 | RPS6 | Down |
| ENSG00000196205 | EEF1A1P5 | Down |
| ENSG00000007264 | MATK | Down |
| ENSG00000212802 | RPL15P3 | Down |
| ENSG00000214194 | SMIM30 | Down |
| ENSG00000165169 | DYNLT3 | Down |
| ENSG00000127184 | COX7C | Down |
| ENSG00000182899 | RPL35A | Down |
| ENSG00000136942 | RPL35 | Down |
| ENSG00000125245 | GPR18 | Down |
| ENSG00000270638 | - | Down |
| ENSG00000233476 | EEF1A1P6 | Down |
| ENSG00000269893 | SNHG8 | Down |
| ENSG00000165512 | ZNF22 | Down |
| ENSG00000150045 | KLRF1 | Down |
| ENSG00000156482 | RPL30 | Down |
| ENSG00000169508 | GPR183 | Down |
| ENSG00000231500 | RPS18 | Down |
| ENSG00000198574 | SH2D1B | Down |
| ENSG00000183918 | SH2D1A | Down |
| ENSG00000169442 | CD52 | Down |
| ENSG00000171858 | RPS21 | Down |
| ENSG00000179841 | AKAP5 | Down |
| ENSG00000198756 | COLGALT2 | Down |
| ENSG00000110848 | CD69 | Down |
| ENSG00000224631 | RPS27AP16 | Down |
| ENSG00000113088 | GZMK | Down |
| ENSG00000139679 | LPAR6 | Down |
| ENSG00000242299 | - | Down |
| ENSG00000177954 | RPS27 | Down |
| ENSG00000162620 | LRRIQ3 | Down |
| ENSG00000145649 | GZMA | Down |
| ENSG00000279377 | - | Down |
| ENSG00000111796 | KLRB1 | Down |
| ENSG00000147604 | RPL7 | Down |

**Supplementary Table 6** The genes identified as SDE with age with severity and the interaction between age and severity included in the model. Positive and negative log2 fold change indicate increasing and decreasing levels with increasing age.

| **ID** | **Gene** | **log2FoldChange** | **B-H p-value** |
| --- | --- | --- | --- |
| ENSG00000242076 | IGKV1-33 | 0.1549 | 2.126E-02 |
| ENSG00000204001 | LCN8 | 0.1706 | 2.126E-02 |
| ENSG00000048052 | HDAC9 | 0.0323 | 2.280E-02 |
| ENSG00000135116 | HRK | 0.1177 | 2.280E-02 |
| ENSG00000272871 | NA | 0.0494 | 2.424E-02 |
| ENSG00000228526 | MIR34AHG | 0.0677 | 2.804E-02 |
| ENSG00000170390 | DCLK2 | 0.1229 | 2.804E-02 |
| ENSG00000124429 | POF1B | 0.0948 | 2.804E-02 |
| ENSG00000102595 | UGGT2 | 0.0410 | 2.804E-02 |
| ENSG00000154262 | ABCA6 | 0.1337 | 2.804E-02 |
| ENSG00000079337 | RAPGEF3 | 0.0893 | 2.927E-02 |
| ENSG00000172869 | DMXL1 | 0.0136 | 3.181E-02 |
| ENSG00000169385 | RNASE2 | 0.0594 | 3.181E-02 |
| ENSG00000100077 | GRK3 | 0.0230 | 3.181E-02 |
| ENSG00000165507 | DEPP1 | 0.1496 | 3.193E-02 |
| ENSG00000104472 | CHRAC1 | -0.0131 | 3.232E-02 |
| ENSG00000196440 | ARMCX4 | 0.0197 | 3.304E-02 |
| ENSG00000156869 | FRRS1 | 0.0308 | 3.993E-02 |
| ENSG00000118432 | CNR1 | 0.1063 | 4.097E-02 |
| ENSG00000198947 | DMD | 0.1304 | 4.097E-02 |
| ENSG00000108950 | FAM20A | 0.1076 | 4.097E-02 |
| ENSG00000210082 | MT-RNR2 | -0.1011 | 4.098E-02 |
| ENSG00000064652 | SNX24 | 0.0243 | 4.388E-02 |
| ENSG00000185710 | SMG1P4 | -0.0801 | 4.388E-02 |
| ENSG00000154258 | ABCA9 | 0.1245 | 4.388E-02 |
